# Clinical utility of Basophil and B-cell biomarkers for monitoring disease activity in food allergy and food oral immunotherapy

**DOI:** 10.1101/2020.01.15.20017541

**Authors:** Theodore Kim, Richard L. Wasserman, Oral Alpan, Atul Shah, Douglas Jones

## Abstract

As research in the field of food allergy is gaining momentum with new and emerging therapies there is need for both researchers and clinicians to have a better understanding on how to put all this new information into context in clinical care. We are continuously learning from other fields, such as oncology, that a one-shoe-fits all type approaches are becoming the practice of the past and there is need to incorporate markers of disease activity as well as drug selection into clinical care. In the United States, this can happen in two ways; laboratory developed testing and companion diagnostics, generally former leading the path to the later. The findings in this letter is a collaboration between four CLIA/CAP accredited in-office flow cytometry laboratories in Utah, New York and Virginia that are part of very busy food allergy clinics directed by board certified Allergist/Immunologists. The identification of changes in basophils and B cells during oral food immunotherapy are proving to be potentially useful markers in monitoring these patients. We show a high ratio of CD63 to CD203c and CD73 expression on B-cells, compared to healthy non-allergic controls and patients who have outgrown their food allergies. This ratio of basophil surface markers as well as B cell CD73 expression drops as patients are undergoing food-OIT. Quite interestingly, we see a similar low pattern in patients who have non-releaser basophils. Altogether these biomarkers are providing useful and important information monitoring patients and we have validated these assays for clinical use as laboratory developed tests. The Basophil Activation Test is used much more routinely outside the United States, and the powerful correlation it provides to oral food challenge outcomes is making it a very attractive tool. We have just submitted two manuscripts on the validation of the BAT as well as sample stability which is under review elsewhere. We expect more utility of the BAT in the United States in the future. Incorporating biomarkers to clinical care of patients with food allergies will provide to be important in assessing efficacy as well as complications of various therapies as well as monitoring the natural resolution of the food allergies.

---

Once peanut allergy develops, the standard of care has been complete avoidance to prevent potentially fatal systemic allergic reactions.. Over the past decade, reports of case series and placebo-controlled studies have shown that the systematic introduction of small amounts of peanut allergen, followed by gradual increases in dose, could prevent or attenuate systemic reactions^1^. A major concern regarding peanut OIT, that the ability to eat the desensitizing allergen will be temporary and lost if regular consumption ceases has not been resolved and remains an important un-answered question in clinical management of these patients. Although the changes in serum peanut specific IgE and IgG4 shows some correlation with OIT outcomes, it is more likely that both serologic and cellular biomarkers will be needed to accurately predict sustained unresponsiveness and/or immune tolerance^2^. Furthermore, the technical validation and performance of such assays in a clinical laboratory is necessary for the assays to be able to guide clinical care. In line of this thought process, we evaluated two candidate assays; a very well established assay that assesses desensitization and a unique B cell assay that may be a marker for change in atopic status over time.

Basophil activation results in the translocation of lysosomal-associated membrane proteins, including CD63, from a predominantly intracellular location to the cell surface. Therefore, CD63 expression measures cells that have undergone degranulation^3^. The ectonucleotide pyrophosphatase/phosphodiesterase (ENPP)-3, CD203c, is upregulated during activation as well. Activation of circulating basophils has been shown to correlate with clinical disease in several contexts including urticaria, anaphylaxis, asthma, food allergy, autoimmune disease and helminth infection^4^.

We studied 34 patients between 2 and 16 years of age receiving peanut oral immunotherapy (peanut-OIT) in this retrospective data analysis. Collection and analysis of data was approved by Western Institutional Review Board (Protocol: 1-824462-1). Peanut allergy was diagnosed by a clinical history of hives, respiratory and/or gastrointestinal symptoms on ingestion of peanut within the 6 months prior to initiating peanut-OIT, a positive skin test and/or peanut specific IgE level measurement (>5kU/L) as well as a basophil activation test to peanut. Of the patients with peanut allergy 35% also had other nut allergies, but no allergies to other foods. Allergic rhinitis and asthma was diagnosed in 90% and 38% respectively. Patients were on different stages of OIT ranging between 2mg dose to maintenance therapy. Male to female ratio was 1:1.2. We also collected data from 19 patients with peanut allergy who are not receiving OIT (ages 2-15), 5 healthy controls (ages 4-16) and 5 subjects (ages 7-18) with near equal M:F ratio, who have outgrown peanut allergy as documented by a low peanut specific IgE titer (<2 kU/L) and passing an oral food challenge. We also included 9 peanut allergic patients with non-releaser basophils (ages 5-14). The non-releaser phenotype can be seen in 15-20% in the general population as well as in food allergic patients and is characterized by the inability to upregulate CD63 on stimulation with anti-IgE as a positive control.

Basophil surface expression of CD63 and CD203c was examined in response to graded concentrations of peanut allergen (Greer Laboratories). The assays were validated in Amerimmune, a CLIA/CAP approved laboratory. Basophils were identified in peripheral blood as CD193^+^CD123^+^IgE^+^ cells. Activated basophils were measured by gating on CD63 positive cells as well as the fold change in CD203c mean fluorescence intensity (MFI) compared to the negative control. Whole blood was stimulated with peanut allergen in concentrations ranging between 10-10,000 ng/ml. Figure 1A shows that the ratio of CD63 to CD203C in peanut-OIT patients is significantly decreased compared to that seen in peanut allergic patients (p < 0.001). The data is from basophils activated with 100 ng/ml concentration of peanut, which showed the best results, although the patterns for the different concentrations were similar. Although there was a decrease in basophil surface CD63 expression in patients undergoing OIT, CD203c expression showed a decrease, no change or slight increase that did not correlate with the stage of peanut OIT, hence the reason we chose to look at ratios rather than individual surface markers. The stage of the peanut-OIT did not correlate with a decrease in the CD63/203c ratio. It is possible that there is a transition phase for the decrease in CD63/CD203c ratio, and a prospective study may provide more evidence for the timing of this finding.

**Figure 1.**
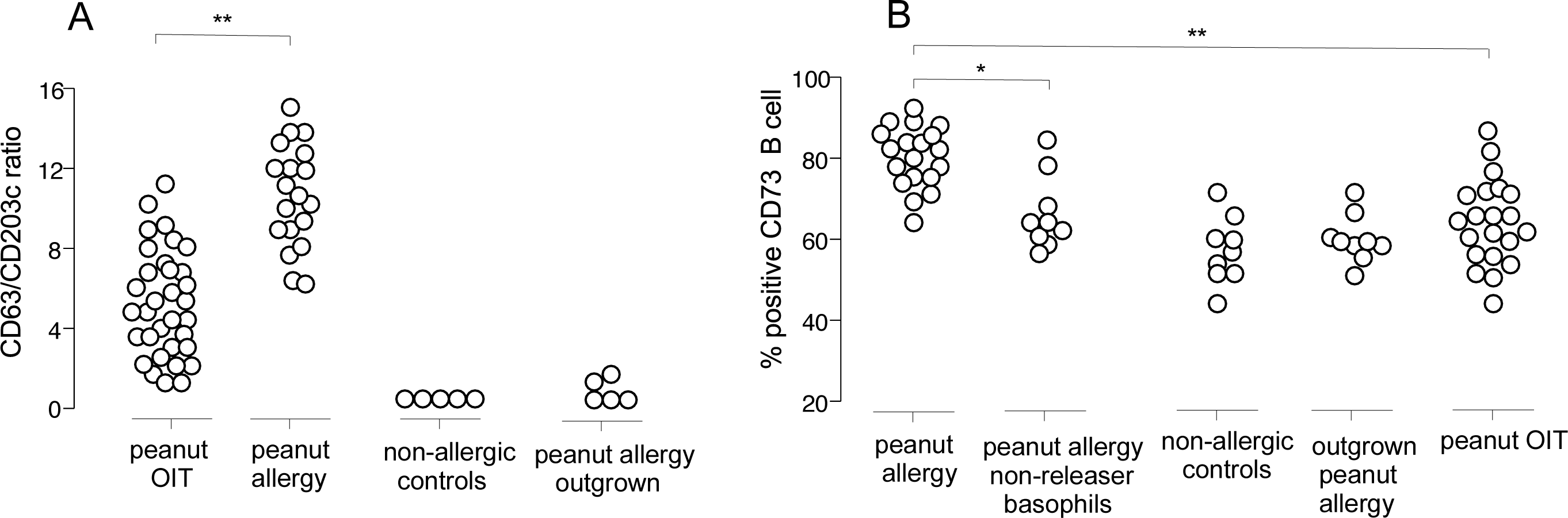
a) CD63/CD203c ratio. CD203c is reported as fold change in mean fluorescence intensity on basophils. CD63 is reported as percent positive basophils that stain positive for CD63. b) CD73 expression on B cells.

We then asked if the changes seen in basophils are accompanied by phenotypic changes in B cells, as one of the goals of OIT is to induce long lasting immunological tolerance. In vitro– activated B cells can downregulate CD73 expression, and inhibit T-cell proliferation and cytokine production^5,6^. Peanut allergic patients showed a significant increase in the expression of CD73, compared to healthy controls or those that have outgrown the peanut allergy, p < 0.001 (Figure 1B). Patients undergoing peanut-OIT also showed a lower CD73 expression, compared to peanut allergic individuals (p < 0.01). In healthy controls, B cell CD73 expression shows an age dependent decrease, and the pattern we are seeing here could be the acceleration of the natural outgrowing process induced by peanut OIT (data not shown). The change in CD73 expression did not correlate with the level of suppression in the CD63/CD203 ratio which suggests that changes in B cells are independent of the basophil desensitization (data not shown).

Immune monitoring during immunotherapy of any kind is essential to better understand success, especially in cases where outcomes are not very obvious during the therapy or the therapy itself leads to complications^7^. We now see this in oncology with checkpoint inhibitors where the utility of this treatment modality is creating a new field in medicine that deals with the complications. At the present time, the status of the peanut allergy is determined by oral food challenges with the attendant risk of anaphylaxis. Clinically available and valuable biomarkers need to be part of management in peanut-OIT.

## Data Availability

No other data available.

